# Pathogenic IgE-recognising epitopes and tolerance-related IgG4 epitopes in patients with and without cow’s milk allergy

**DOI:** 10.1101/2025.03.13.25323876

**Authors:** Kiyoaki Ito, Ikuo Okafuji, Mitsuhiro Kawano, Yusei Ohshima, Yoshihiro Watanabe

**Affiliations:** IgG4-related Immunology, Graduate School of Medical Science, Kanazawa University. 13-1 Takara-machi, Kanazawa 920-8641, Ishikawa, Japan; Rheumatology of Hospital, Kanazawa University. Kanazawa University. 13-1 Takara-machi, Kanazawa 920-8641, Ishikawa, Japan; Pediatrics, Kobe City Medical Center, Citizen’s General Hospital, 2-1-1 Minatojima Minami-cho, Chuo-ku, Kobe 650-0047, Hyogo, Japan; Hematology, Immunology and Internal Medicine of Hospital, Kanazawa Medical University. 1-1 Daigaku, Uchinada-cho, Kahoku0gun 920-0293, Ihikawa, Japan; Pediatrics of Medicine, Fukui University. 3-3 Simoaitsuki 2, Matsuoka, Eiheiji-cho, Yoshida-gun 910-1193, Fukui, Japan

**Keywords:** IgE, IgG4, epitope profile, cow’s milk allergy, 14 epitopes-ALL, pathogenic, tolerance

## Abstract

**Introduction:** An enzyme-linked immunosorbent assay (ELISA) system capable of profiling IgE-recognising epitopes in patients with cow’s milk allergy (CMA) was established previously. This assay can reveal qualitative differences and imbalances between IgE- and IgG4-recognising epitopes as well as quantify IgE specific to the mixture of 14 epitopes (14 epitopes-ALL) that correlate with the value obtained using the ImmunoCAP sIgE(milk) test. To apply this system clinically, we determined the cutoff range of IgE contents detected by the 14 epitopes-ALL, and identified pathogenic IgE-epitopes and tolerance-related IgG4-epitopes by comparing the profiles of both IgE and IgG4.

**Methods:** Serum samples from 38 patients with CMA and 34 non-CMA volunteers were assessed to determine their IgE levels towards 14 epitopes using our ELISA system. Epitope profiles of the samples were analysed individually. Using clinical data on oral immunotherapy status and allowed milk intake assessed by the oral food challenge test, the IgE and IgG4 profiles were compared to extract pathogenic, non-pathogenic, and tolerance-related epitope candidates.

**Results:** The cutoff range of IgE was 1.5–4.5 ng/mL with an area under the curve of 89% in receiver operating characteristic analysis. Serum samples of ImmunoCAP class 3 and lower classes were divided into upper and lower proportions by this cutoff range, which can be useful for predicting risk of eliciting symptoms by allergenic food exposure. Extraction of candidate pathogenic and non-pathogenic epitopes showed that pathogenic epitopes formed a cluster in hydrophobic regions of caseins and appeared to be near each other on the molten globule of micelles.

**Conclusions:** The 14 epitopes of cow’s milk allergens are useful for determining the allergen-specific IgE concentration in patients with CMA. The IgE vs. IgG4 profile identified pathogenic, non-pathogenic, and tolerance-related epitopes. This profiling analysis may explain why oral immunotherapy is effective in some individuals but not others.

**Key Messages:**

- IgE antibodies recognising 14 cow’s milk allergen epitopes correlate with IgE values detected using ImmunoCAP.
- This test can accurately assess risk of symptoms in patients with class 3 ImmunoCAP sIgE.
- Epitope-based quantitative/qualitative analyses of antibodies are reliable for assessing allergic symptoms and oral immunotherapy effects.

## Introduction

In patients with food allergies, oral immunotherapy (OIT) and diet therapy containing asymptomatic amounts of allergenic foods can lead to desensitisation or sustained unresponsiveness to the allergen. However, in some cases, allergenic foods must be avoided completely. Unresponsiveness can be addressed through controlled intervals of exposure to allergens that are not well-tolerated as part of the desensitisation process. Although the risk of anaphylaxis must be carefully considered in patients who do not respond to OIT with allergenic foods, its onset is difficult to predict [1, 2]. Allergen food or component-specific IgE values measured using the ImmunoCAP test and a skin prick test are currently used as predictive biomarkers [3, 4]. Both tests can assess the allergic status but exhibit uncertainty in predicting the risk of severe allergic symptoms associated with accidental intake of allergen foods. For patients of ImmunoCAP-sIgE (milk) class 3, the probability of allergic reactions varies from 30% to 70%. Therefore, the oral allergen food challenge (OFC) test is conducted to directly assess symptom induction caused by allergenic food intake to set a threshold level; however, this test is time-consuming and may induce anaphylaxis [5–7].

OIT can induce ‘blocking antibodies (Abs)’ such as IgG4 that are specific to allergen foods. Studies have attempted to determine how allergen-specific IgG4 suppresses IgE responses. However, the detailed mechanism of blocking IgG4 Abs in ‘effective’ and ‘ineffective’ cases remains unclear. Allergen-specific or component-specific IgG4 Ab levels and the IgG4/IgE ratio may be useful biomarkers indicating the efficacy of OIT treatment [8, 9]. However, there is no correlation between the IgG4/IgE ratio and amount of allergen food consumed [10–13]. Thus, whether allergen-specific IgG4 values can be used as biomarkers in clinical settings remains unclear [8, 14].

IgG4 is a unique IgG subclass of Abs that does not have effector functions and is induced following a series of Th2-type and related regulatory immune responses. IgG4 Abs are thought to negatively regulate IgE-allergen binding, followed by mast cell/basophile activation via FcεRI crosslinking, enhancement of allergen binding via FcεRII, and other IgE functions [15, 16]. However, IgE-secreting B cells and IgG4- producing B cells exhibit differences in allergen recognition because differentiation into either IgE+ B cells or IgG4+ B cells is separately induced by allergen-specific IgG1 memory B cells. In fact, the peptide epitopes recognised by either IgE or IgG4 differed according to peptide array, phage display, and single-cell level Ig-sequence analysis of their complementarity-determining regions [12, 13, 17–19]. That is, mismatch occurs between the IgE- and IgG4-recognising epitopes, and increasing the amount of IgG4 Ab does not directly lead to blocking of IgE-mediated responses [11, 20, 21].

We recently reported that the 14 epitopes in milk allergens are useful as a reliable and simple epitope profiling system, and that the IgE contents detected by the 14 epitopes correlate with ImmunoCAP-sIgE (milk) values in patients with cow’s milk allergy (CMA) [22]. This enzyme-linked immunosorbent assay (ELISA)-based profiling method and Luminex system, as another reliable profiling method, can clarify epitope profiles recognised by IgE and other classes of Abs [20, 22, 23]. Using the defined 14 epitopes, we analysed IgE and IgG4 profiles in serum samples from patients who received OIT of cow’s milk or diet therapy with an amount of milk intake that did not induce symptoms. We examined whether this profiling analysis can reveal qualitative differences between the epitopes recognised by IgE versus IgG4 in patient serum. The pathogenic IgE epitopes related to the severity of allergenic symptoms and IgG4 epitopes related to sustained unresponsiveness and tolerance were determined. The contribution of IgG4 to inducing and maintaining remission or tolerance in OIT responders and nonallergic participants was examined.

## 2. Methods

### 2-1. Samples, data collection, and ethics statement

Patients undergoing OIT for CMA at the University of Fukui Hospital and Kobe City Medical Center General Hospital participated in the study (No. 20210138). The study was approved by the University of Fukui Ethics Committee. Non-CMA volunteers at Kanazawa University Hospital also participated in this study (No. 2024-148, No. 2024- 131), which was approved by the Kanazawa University Ethics Committee. Informed consent was obtained in accordance with the principles of the Declaration of Helsinki. Serum samples from individual participants were collected and stored at −80°C until use.

### 2-2. Materials and reagents

The secondary Abs of horseradish peroxidase-conjugated anti-human IgE (MCA E-09-HRP) and human IgG4 (MCA DG-01-HRP) were purchased from Yamasa (Tokyo, Japan). The peptide epitopes and reagents for ELISA are detailed in our previous report [22].

### 2-3. ELISA and calculation of Ab contents (IgE and IgG4) specific to defined epitopes

Microtiter plates (Maxisorp; Thermo Fisher Scientific, Waltham, MA, USA) were coated with streptavidin at 100 ng/well (epitope profiling) or 300 ng/well (14 epitopes-ALL) in 50 μL phosphate-buffered saline. The detailed procedures including calculation of IgE and IgG4 levels using standard recombinant Abs have been described previously [22].

### 2-4. Profiling analysis of IgE- vs. IgG4-recognising epitopes

At appropriate dilutions determined by calculating the IgE and IgG4 contents specific to the 14 epitopes-ALL, the epitope profiles were obtained in IgE and IgG4 using microtiter plates coated with each epitope. The IgE and IgG4 profiles were compared using clinical information such as the results of ImmunoCAP-sIgE (milk) test, the total IgE value, and allowed volume of milk intake by the OFC test, in addition to the concentrations of IgE and IgG4 Abs specific to the 14 epitopes-ALL and the IgG4/IgE ratio.

## 3. RESULTS

### 3-1. Provisional cutoff range of IgE specific to 14 epitopes-ALL using serum samples from non-CMA volunteers

Using 14 individual epitopes, we analysed the IgE profiles of patients with CMA and non-allergic volunteers. Fig. 1A shows an example profile. The serum from patient-2906, which was categorised as class 3 by ImmunoCAP-sIgE (milk), contained IgE Abs specific to epitope No. 5. E, itope Nos. 3–6 and 10 were secondarily detected by IgE, resulting in accumulation of IgE Abs in the 14 epitopes-ALL (red dotted box). The IgE content of patient-2906 was 4.6 ng/mL based on comparison with a standard curve [22]. Non-CMA volunteer A446 showed no significantly positive epitopes. The total amount of IgE specific to the 14 epitopes-ALL was 0.56 ng/mL.

**FIGURE 1.**
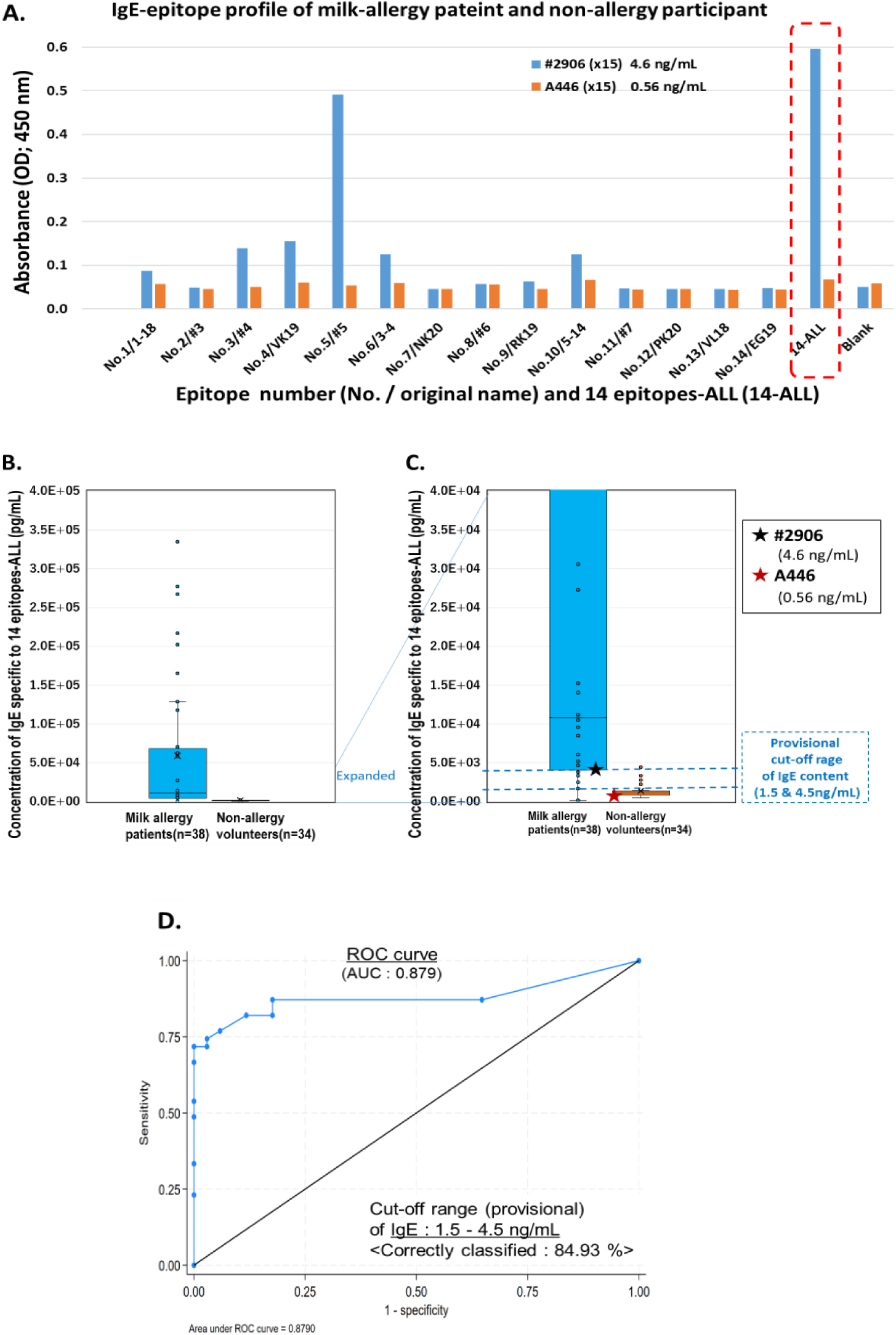
Epitope profiling of serum from patients with cow’s milk allergy (CMA) and non-allergic volunteers. **A.** Individual serum with appropriate dilution was assessed to determine its recognition towards 14 individual epitopes and epitopes-ALL. Representative profile data of two serum samples from patient with CMA (2906) and non-allergic volunteer (A446) are depicted as a bar graph. Representative data of mean values in duplicate measurement are shown in multiple experiments. **B and C.** IgE contents recognising epitopes-ALL of all patients with CMA and non-allergic volunteers were plotted as a bar graph with provisional cutoff range (1.5 and 4.5 ng/mL). **D.** Receiver operating characteristic curve of IgE contents between patients with CMA and non-CMA volunteers. Area under the curve (AUC) is 0.879, and the cutoff range is 1.5 to 4.5 ng/mL, which correctly classified 84.93% of samples.

Serum samples from 32 non-allergic volunteers were assessed to determine whether IgE is specific to the 14 epitopes. In Figs. 1B and 1C, the IgE levels in serum samples of non-allergic volunteers are plotted for 38 patients with CMA. Using provisional cutoff values of 4.5 and 1.5 ng/mL, all non-allergic volunteers were considered completely negative and more than 83% were considered negative, respectively. In Fig. 1C, the IgE contents in representative serum samples of patient-2906 with CMA and non-allergic volunteer A446 were plotted as ★ marks (black and red, respectively). In Fig. 1D, the receiver operating characteristic (ROC) curve for each criterion showed that the provisional high (4.5 ng/mL) and low (1.5 ng/mL) concentrations led to correctly classified results in 84.9% of cases. The area under the ROC curve was 87.9%. The results of positivity, sensitivity, and specificity analyses are shown in Supplementary Table 1 and 2.

### 3-2. Provisional cutoff range divided serum samples of patients with ImmunoCAP-sIgE (milk) class 3 into two populations

We previously determined the correlation in plots between IgE contents of 14 epitopes-ALL vs. ImmunoCAP sIgE (milk) [22]. Fig. 2A shows the correlation plots with the upper and lower contents of the provisional cutoff range (dotted grey lines; 4.5 and 1.5 ng/mL). No serum samples were below the criteria in patients categorised as class 4– 6 detected using ImmunoCAP-sIgE (milk). Additionally, almost all samples detected as ImmunoCAP class 2 and 1 were below the criteria, whereas at least one sample exceeded the high value of the provisional cutoff range.

**FIGURE 2.**
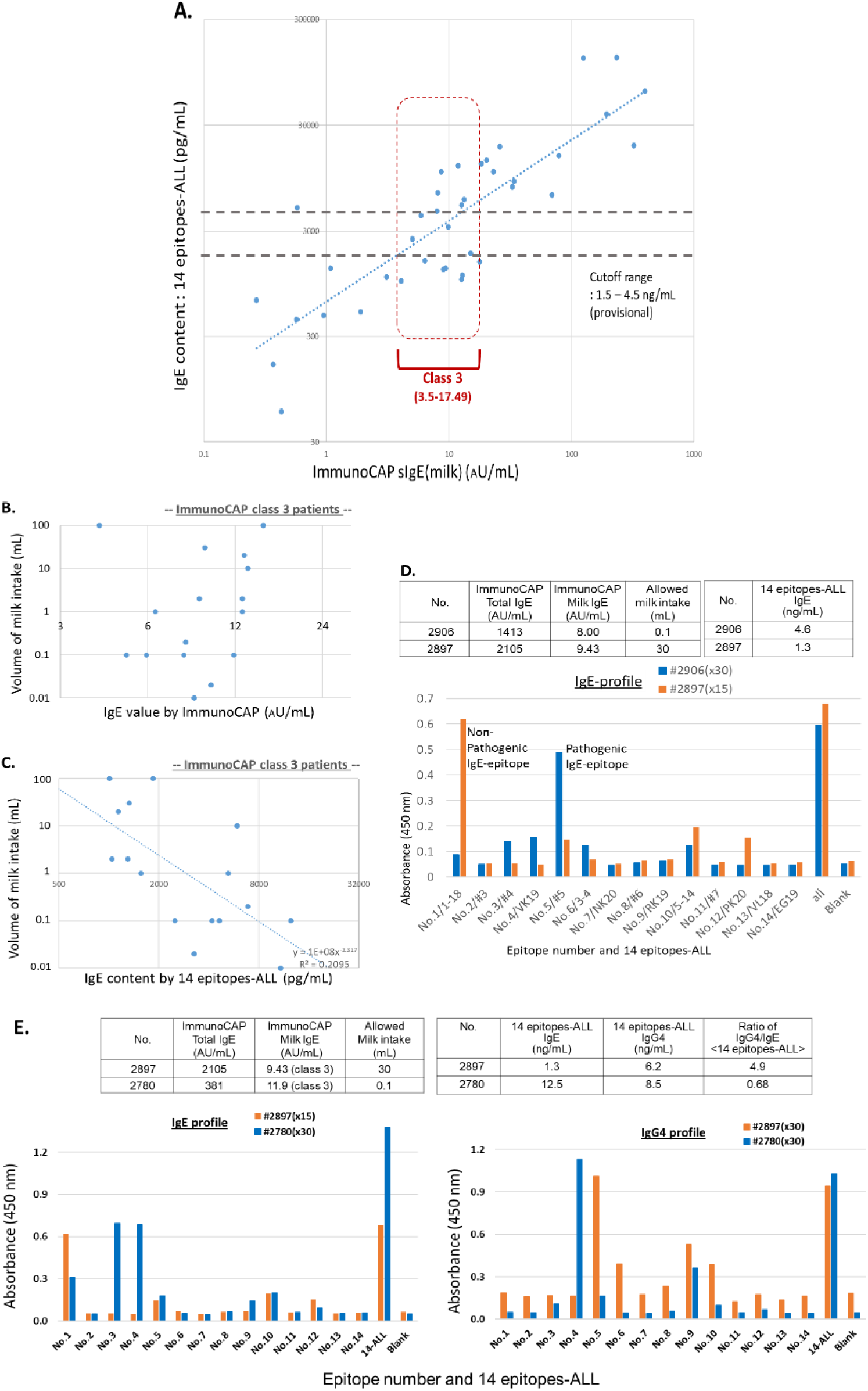
Cutoff range divides ImmunoCAP class 3 samples into two populations. **A**. IgE contents recognising the 14 epitopes-ALL in all patients with CMA were plotted with ImmunoCAP sIgE (milk) values. The provisional cutoff range of vertical axis is shown as dotted lines. **B and C.** Serum samples from patients with ImmunoCAP sIgE class 3 were further analysed in terms of volume of milk intake assessed by oral food challenge (OFC) test in reference to the IgE values detected by ImmunoCAP-sIgE (milk) (**B)** or IgE contents detected by 14 epitopes-ALL (**C**). **D.** IgE profiles of representative two samples of ImmunoCAP class 3. The profiles were further evaluated using clinical data of ImmunoCAP IgE values, allowed milk volume by OFC test, and IgE contents specific to 14 epitopes-ALL listed in the upper tables. Major epitopes detected by IgE were assigned as “non-pathogenic” or “pathogenic” epitope candidates. **E.** Representative profiles of IgE (left panel) and IgG4 (right panel) with clinical data (upper left table) and calculated Abs data obtained by 14 epitopes-ALL (upper right table) are shown. Major epitopes detected in the profiles of IgE and IgG4 were determined as the criteria within x1/3 values of OD (optical density) from the highest OD value of recognising epitope. These are counted and described in Table 1 as frequencies of positive samples. The comparative analysis and explanation of pathogenic vs. non-pathogenic epitopes were described in the Results section.

**Table 1.**
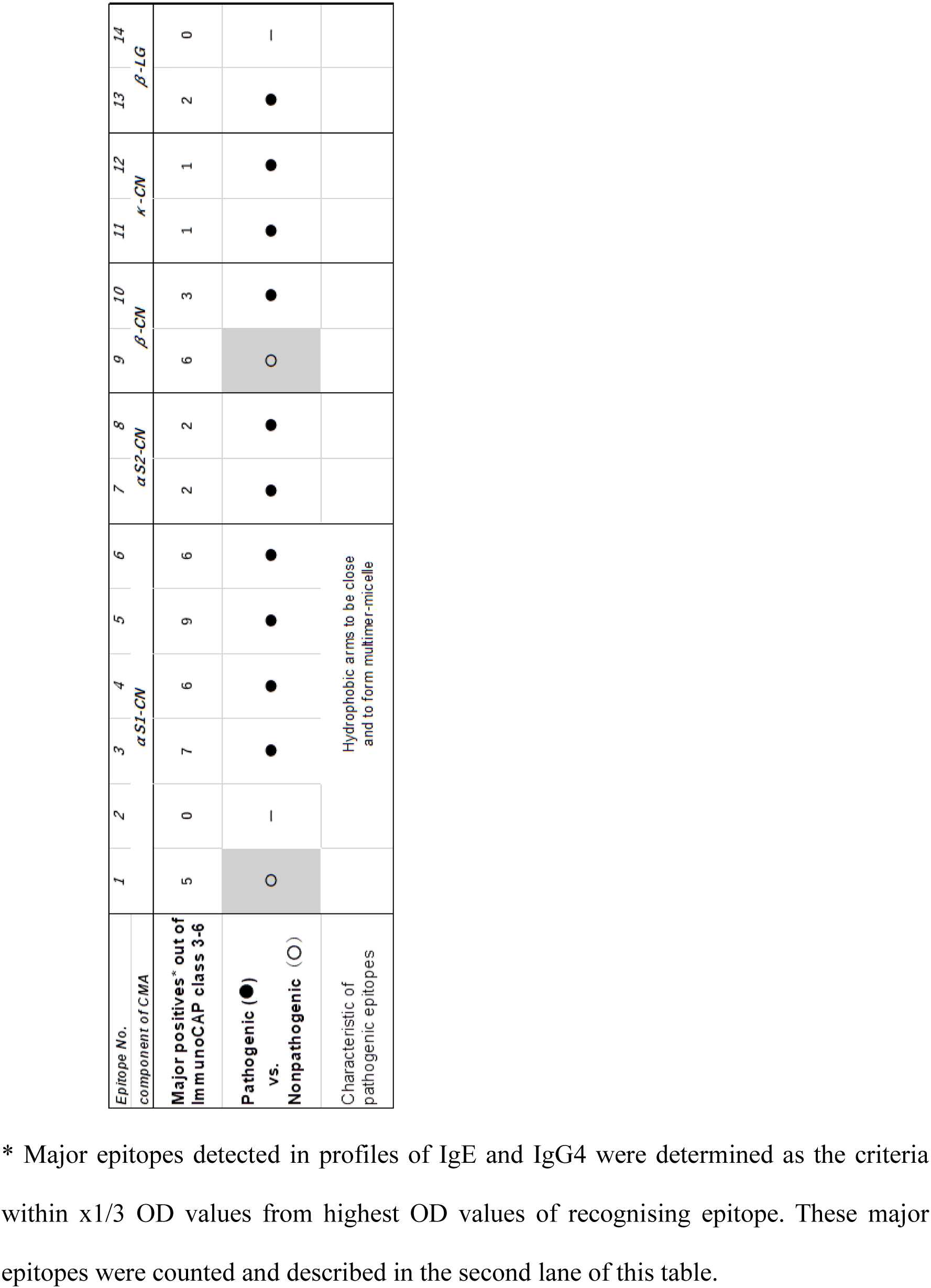
Identification of ‘pathogenic’ and ‘non-pathogenic’ IgE epitope candidates.

In class 3 serum samples detected using ImmunoCAP, the IgE contents detected by 14 epitopes-ALL varied above and below the provisional cutoff criteria. This variation may be related to the probability of eliciting allergic symptoms in ImmunoCAP class 3 patients, which is 30–70% following allergen exposure [6, 7]. These results indicate that IgE quantification detected by 14 epitopes-ALL can divide patients classified as ImmunoCAP class 3 into two populations with high and low risks of inducing allergic symptoms following milk intake.

To confirm this hypothesis, we examined the correlation between the volume of milk intake assessed by the OFC test and IgE content measured by the 14 epitopes-ALL, focusing on ImmunoCAP class 3 patients. Milk intake was not related to ImmunoCAP IgE levels (Fig. 2B). In contrast, Fig. 2C shows a tendency of inverse correlation between vertical and horizontal axes. This result indicates that ImmunoCAP class 3 patients who were unable to drink cow’s milk had high IgE levels detected by 14 epitopes-ALL and that IgE levels specific to14 epitopes-ALL decreased with consumption of increasing milk volumes.

The results in patients with ImmunoCAP class 3 suggest that the defined 14 epitopes can reveal pathogenic IgE epitopes. We further examined the epitope profiles in serum samples from all patients. Fig. 2D shows two representative serum samples with clinical data such as ImmunoCAP sIgE (milk) and allowed volume of milk intake determined using the OFC test. Although both serum samples were categorised as ImmunoCAP class 3, the epitope profiles largely differed from each other. Based on clinical data, we assigned the No. 5 epitope as a candidate pathogenic IgE-epitope and No. 1 epitope as a candidate non-pathogenic IgE-epitope, as patient-2906 could not drink more than 0.1 mL milk but patient-2897 could drink 30 mL of milk without eliciting allergic symptoms. As shown in the upper right table of Fig. 2D, the contents of IgE detected by the 14 epitopes-ALL were calculated. The IgE content showed that patient-2906 exceeded the provisional cutoff criteria and that patient-2897 was below the cutoff criteria.

### 3-3. Comparative analysis of IgE profile vs IgG4 profile

As shown in Fig. 2D, the epitope profile of IgE can identify pathogenic and non-pathogenic epitopes. Furthermore, comparison of the IgG4 to the IgE profile revealed the balance between pathogenic IgE versus blocking IgG4. It is thought that IgG4 Abs specific to allergens compete with IgE at the epitope level in OIT-effective patients. To determine the balance of each IgE versus IgG4, we compared each profile.

In Fig. 2E, two IgE profiles and two IgG4 profiles are shown with clinical and measurement data using the 14 epitopes-ALL. One case was OIT-effective (patient-2897) and another was OIT-ineffective (patient-2780); both serum samples were categorised as ImmunoCAP class 3. In OIT-ineffective patient-2780, IgE Abs specific to epitopes No. 3 and No. 4 localised in αS1 casein were major epitopes and were very likely pathogenic. From both IgE profiles of OIT-ineffective patients, epitope Nos. 3–5 were defined as pathogenic. In contrast, the IgG4 profile of patient-2780 differed from the IgE profile, and no IgG4 Abs recognised epitope No. 3 compared with the large amount of No.4 epitope-specific IgG4. These data indicate that patient-2897 has different IgE and IgG4 qualities in terms of epitope specificity towards individual epitopes. The profile set of IgE versus IgG4 of patient-2897 showed a major IgG4 Abs-specific to the No.5 epitope as a pathogenic epitope. However, there was no No. 5 epitope-specific IgE, and the No. 1 epitope-specific IgE was non-pathogenic.

Both patients showed similar ImmunoCAP IgE values with class 3, but the IgE contents detected by 14 epitopes-ALL largely differed from each other. Patient-2897 was below the cutoff criteria and patient-2780 exceeded the cutoff value. According to profile analysis, patient-2897 sustained IgG4 Abs specific to the No. 5 epitope without the appearance of IgE recognising pathogenic epitopes Nos. 3 and 5. In patient-2780, IgE Abs specific to Nos. 3 and 4 epitopes were detected, but No. 3 epitope-specific IgG4 Abs were not observed. The profiles were clearly imbalanced each other according to comparative analysis.

### 3-4. Extraction of pathogenic and non-pathogenic IgE epitope candidates and risk of eliciting allergic symptoms

Comparative analysis of IgE and IgG4 with clinical data for all serum samples (Fig. 2E, Supplementary S-Fig. 1) revealed candidates of pathogenic and non-pathogenic IgE epitopes. These candidates are summarised in Table 1. Importantly, there were at least two epitopes of non-pathogenic IgE. As similar to the profile of patient-2897, IgE profiles were positive for the No.1 epitope in some patients who consumed a sufficient volume of milk in diet therapy (Supplementary S-Fig. 1). Epitope No. 1 is in αS1-casein and epitope No. 9 is in β-casein, and both are localised in N-terminal regions [24]. Furthermore, a cluster of pathogenic IgE epitope Nos. 3–6 localise in the hydrophobic regions of α-casein with high frequencies [24–26].

Based on these findings, we evaluated the risk of inducing anaphylaxis. An IgE level exceeding the cutoff criteria is critical for defining high-risk populations. Comparative analysis of the IgE versus IgG4 profiles based on “pathogenic” and “nonpathogenic” epitopes is important for determining the risk of eliciting allergic symptoms in patients below the cutoff. This comparative analysis of profile may also be useful for explaining the effectiveness and ineffectiveness of OIT and OFC-based diet therapy.

### 3-5. Discovery of IgG4-positive to 14 epitopes-ALL in non-allergic volunteers and breakdown into individual epitope recognition

As shown in Fig. 1, non-CMA volunteers were considered as negative for IgE specific to the 14 epitopes-ALL. We tested the presence of IgG4-positive results in the sera of non-CMA volunteers and identified some positive sera at very high levels in IgG4 specific to the 14 epitopes-ALL. Fig. 3A shows representative data, and two to four serum samples were positive among the 12 samples. We next examined the four serum samples (A520/A631 and A456/A659) to determine the epitope-profile of the 14 individual epitopes. Two samples showed very strong signals for detecting single epitope No. 5 recognized by IgG4, and the other two samples (A456/A659) were positive for IgG4, detecting multiple epitopes Nos. 4 and 7 at similar levels (Fig. 3B).

**FIGURE 3.**
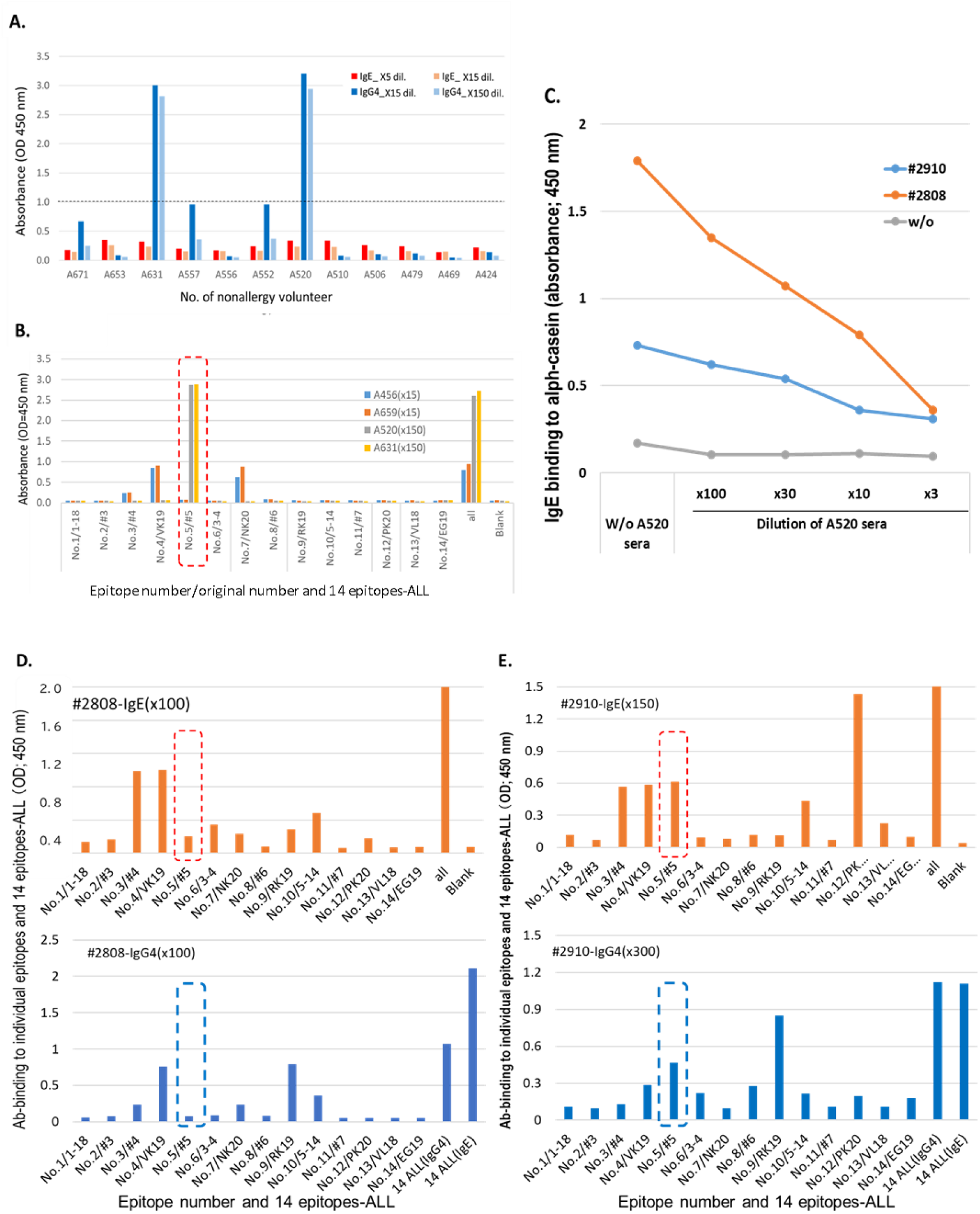
IgG4-positive results whose serum samples recognise 14 epitopes-ALL and few defined epitopes. **A.** Representative bar graph for screening IgG4-positive results in serum samples of non-CMA volunteers are shown with their dilutions described. The 14 epitopes-ALL were used to detect IgE and IgG4 Abs. **B.** Epitope profile as breakdown analysis of IgG4-positive results with appropriate dilutions of samples. **C.** Competition analysis of IgG4-positive serum (A520) on IgE-binding to α-casein of patient-2808 with CMA and patient-2910. **D and E.** Epitope profiles of IgE and IgG4 of patient-2808 with CMA and patient-2910. These profiles showed that No. 5 epitope recognised by IgE and IgG4 Abs is not a major epitope in patient-2808 and one of major epitopes in patient-2910.

These epitopes recognized by IgG4 were categorised as pathogenic IgE epitopes, such as Nos. 5, 4, and 7 (Table 1). These samples of two sets were collected at 2–3-month intervals, so that the IgG4 intensity specific to milk allergen epitopes was maintained at a similar level during these periods. These results indicate that a large population of non-allergic individuals maintain IgG4 Abs specific to the defined epitopes. Interestingly, IgG4 responses in non-allergic individuals may already be skewed to a single or few specific epitopes. We had evaluated which OIT-effective patient serum inhibited IgE binding to α-casein in pooled sera from seven OIT-ineffective patients (Supplementary S- Fig. 2). Non-CMA volunteer serum was added in the α-casein-binding assay of pooled sera, in which the IgE profile was clarified for the 14 individual epitopes. Fig. 3C shows the inhibition by A631 serum, which contained only No. 5 epitope-specific IgG4 but no IgE specific to α-casein, on IgE binding to α-casein of serum samples in allergy patient-2901 and −2808 who were resistant to OIT treatment with only 0.1 mL of milk intake.

As shown in Figs. 3D and 3E, the IgE and IgG4 profiles of patient-2901 and patient-2808 differed, although both patients were resistant to OIT. Addition of A520/A631 volunteer serum inhibited this IgE-binding. Furthermore, the inhibitory efficacy may be dependent on IgE profiles focusing on a cluster of αS1-casein epitopes (patient-2808) or spread to other component epitopes of κ-casein (patient-2910). These results demonstrate that No. 5 epitope-recognising IgG4 Abs can inhibit IgE binding to αS1-casein of sera from patients with CMA who are resistant to OIT.

## 4. Discussion

In previous reports and in this study, each IgE profile differed among individuals, and IgE versus IgG4 profiles differed in the same individual (Fig. 2 and Supplementary S-Fig. 1) [11, 12, 21, 27–29]. This difference is derived from independent switching of IgE- or IgG4-producing B cells from IgG1 memory B cells upon allergen exposure or OIT treatment [17, 30, 31]. This difference in profile and mismatch of pathogenic IgE epitopes versus epitopes recognised by IgG4 Abs made it difficult to predict the risk of inducing severe allergic reactions. IgE titres and ratios of IgE/other Ig classes specific to allergen proteins cannot accurately predict the risk of severe allergy in ImmunoCAP class 3 patients, as current diagnostic systems detect IgE values including non-pathogenic IgE and detect IgG4 that recognises epitopes unrelated to inhibition of pathogenic IgE responses (also refer to S-Fig. 3). Pathogenic epitopes were detected as clusters inside allergen αS1-casein (Table 1). We also found that two IgE epitopes of the N-terminal parts of αS1-casein and β-casein were frequently detected as non-pathogenic epitopes in patients with allergy at occasionally high intensities (Fig. 2D, Supplementary S-Fig. 1A). In fact, patients who were able to drink a large volume of milk following OIT treatment still showed a high IgE titre towards the N-terminal parts of allergens. Epitope profiling analysis of IgE Abs with reference to the volume of milk intake tested by OFC identified non-pathogenic IgE epitopes among the 14 epitopes. In contrast, pathogenic IgE epitopes were identified in serum samples derived from patients who did not respond to OIT or who could not increase milk intake over 0.1 mL without allergic reactions.

Table 1 lists both pathogenic and non-pathogenic epitopes recognised by IgE. Namely, epitopes Nos. 3–6 are pathogenic epitopes of αS1-casein that form a cluster of epitopes, and some pathogenic epitopes in αS2-casein, β-casein, κ-casein, and β-lactoglobulin were determined. We also identified two non-pathogenic epitopes localised in the N-terminal regions of αS1-casein and β-casein (Table 1). Milk casein forms a molten globular structure; the N-terminal regions are likely flexible and unstable or can be easily hydrolysed as soluble fragments [24–26]. Based on these biophysical characteristics, IgE binding to these regions could not involve the formation of a multimer complex. In contrast, epitopes of a cluster in αS1-casein were nearby and formed multimer complexes with IgE bound to FcεRI on basophils, mast cells, and other effector cells [32, 33]. The ability to block IgE responses may occur through IgG4-recognising pathogenic IgE epitopes. Furthermore, IgG4 Abs that recognize epitopes identical and nearby pathogenic IgE-epitopes can likely block IgE-responses.

Interestingly, some non-allergic and healthy individuals with neither milk allergy nor specific IgE to allergens were positive for IgG4, recognising 14 epitopes (Fig. 3). These findings indicate a mechanism for maintaining tolerance-related IgG4 Abs. In the case of T cell immunity, three major mechanisms of antigen-specific tolerance have been demonstrated: (i) clonal deletion, (ii) clonal anergy, and (iii) active suppression/immune deviation. In the third mechanism of active suppression/immune deviation, Treg cells are highly involved in cell-cell interactions and regulatory cytokine production [34, 35]. B cells with Treg-help produce allergen-specific IgG4 Abs to inhibit pathogenic IgE reactions [16, 36]. Therefore, IgG4 likely functions in mechanism (iii) in B cell tolerance. Thus, certain IgG4 supplements can broadly inhibit IgE responses in an allergen-specific manner, and the allergen-IgG4 complex can contribute to tolerance maintenance/inducing mechanisms. IgG4-producing B cells are thought to have a memory pool [17, 37]. Once memory IgG4+ B cells are produced and established, allergen tolerance may be maintained throughout life upon frequent or sometimes activation of memory lymphocytes. We found that the same epitope-recognising IgG4 Abs existed in non-allergic volunteers and were induced in OIT-effective patients, supporting this hypothesis (Fig. 3B and Supplementary S-Fig. 2A). We also identified some non-allergic volunteers with No. 5 epitope-specific IgG4 in significant frequency (to be published elsewhere).

IgG4 elevation is accomplished by IL-10, IL-35, TGFβ, and other cytokines produced by Tregs and Bregs [34, 35]. As a result, class switching from IgG1 to IgG4 is achieved and leading to allergen tolerance. IgG4 Abs specific to certain epitopes have a wide coverage range for IgE inhibition. For example, the No. 5 epitope-specific IgG4 Ab inhibited the nearby epitope-recognising IgE specific to α-casein (Fig. 3C). For cat allergy, administration of two IgG4 Abs to patients improved visual analogue scale scores, suggesting that a few epitopes-specific Abs can maintain and/or induce unresponsiveness [38, 39]. Our data (Fig. 4B) also showed that one or two key epitope(s) recognising IgG4 Abs are sufficient to induce or maintain the effectiveness of OIT and diet therapy. In theory, IgG4 Abs can extend their suppressive functions via hetero bi-specific IgG4 Abs, which can recognise different allergen components and engage the FcγRIIb inhibitory IgG receptor in a dual allergen-specific manner [32, 36]. The defined epitope recognised numerous IgG4 Abs in non-allergic subjects, which is consistent with the pathogenic epitopes observed in many patients with CMA. This observation suggests that these IgG4 Abs contribute to the ‘remission/tolerance maintenance’ mechanism. Epitope analysis in a wide range of non-allergic subjects may reveal the major tolerance-inducing IgG4 Abs. No. 5 epitope-specific IgG4 is one such candidate, and these IgG4 may induce unresponsiveness and tolerance in patients with severe CMA.

Regarding limitation of this study, the cutoff value of 14 epitopes-ALL detection was set as the provisional range. Based on the results in this study, cutoff value should be fixed with following studies. The usefulness and advantage of 14 epitopes-ALL detection have to be confirmed with enough number of patients with ImmunoCAP class 3 since we could not focus on this category patients at the setting of this study. And, pathogenic and non-pathogenic epitopes of IgE Abs have to be confirmed with prospective study setting.

## 5. Conclusion

IgE and IgG4 Abs were quantitatively and qualitatively evaluated in patients with CMA and non-CMA volunteers using an ELISA system of 14 defined epitopes. This ELISA system can be applied in the clinic as a better diagnostic for CMA and OIT treatments to accurately predict the risk of eliciting allergic symptoms.

## Data Availability

All data produced in the present study are available upon reasonable request to the authors

https://www.sciencedirect.com/science/article/pii/S0022175924001583?via%3Dihub

## Data Availability Statement

Expects Data Sharing

## IRB Statement

The approved clinical researches for this study were detailed in Methods section.

## Acknowledgments

We thank Dr. Shigeki Kato for providing standard proteins of cow’s milk allergens and original mouse mAbs to recognise α−casein or β−lactoglobulin. We also thank Satoko Tamai, Natsuko Ohbayashi, and Takako Ohbayashi for performing measurements of IgE contents specific to 14 epitopes-ALL and IgE vs. IgG4 profiles towards 14 individual epitopes. For preparing this manuscript, we would like to thank Editage (www.editage.jp) for English language editing.

## Funding

This study was funded by JSPS KAKENHI (Grant Number JP 23K07892) and AMED (Grant Number 20ek0410070h0001 and 23yrn0l26802j0002) and supported the by JST Program for co-creating startup ecosystem (Grant Number JPMJSF2318, Japan).

## Author contributions

KI and YW conducted ELISA measurement and wrote manuscript. IO and YO prepared clinical samples and data of allergy patients, MK prepared serum samples of non-allergy patients and suggested contents of this study. YO and YW designed outline of this study and conducted it thoroughly, then made discussion in terms of significance of this study.

## Supporting Information

### 1) Determination of cutoff range between CMA patients and non-CMA volunteers

ROC analysis was done with classified table of both populations in terms of IgE contents detected by 14 epitopes-ALL EKISA system. The classified table was shown in Supplementary Table 1, and the result by ROC analysis was shown in Supplementary Table 2 and Fig. 1C.

**Supplementary Table 1. A.** <u>ROC analysis of IgE contents in serum samples of between patient with CMA and non-CMA volunteers</u>. IgE contents were measured with 14 epitopes-ALL detection system and classified in the left column of table. The numbers of serum samples classified were counted in each population of patients and volunteers. **B.** These classified numbers in each lane were evaluated with ROC analysis. The values of “correctly classified” was highest at the range of 1.5 ng/mL to 4.5 ng/mL, and AUC value of this ROC curve was 88.0%.

**Supplementary Table 1.** Classification of IgE contents specific to the 14 epitopes-ALL in CMA patients and non-CMA volunteers.

| Data value<br>(ng/mL) | patients | non-patients | maker |
| --- | --- | --- | --- |
| <1.0 | 5 | 12 | 1 |
| 1.0 <1.5 | 0 | 16 | 2 |
| 1.5 <2.0 | 2 | 0 | 3 |
| 2.0 <2.5 | 0 | 2 | 4 |
| 2.5 <3.0 | 2 | 2 | 5 |
| 3.0 <3.5 | 1 | 1 | 6 |
| 3.5 <4.0 | 1 | 0 | 7 |
| 4.0 <4.5 | 0 | 1 | 8 |
| 4.5 <5.0 | 2 | 0 | 9 |
| 5.0 <7.0 | 5 | 0 | 10 |
| 7.0 <10.0 | 2 | 0 | 11 |
| 10 <30 | 6 | 0 | 12 |
| 30 <100 | 4 | 0 | 13 |
| 100 <1000 | 9 | 0 | 14 |

**Supplementary Table 2.** Sensitivity and Specificity of ROC analysis.

| Detailed report of sensitivity and specificity |  |  |  |  |  |
| --- | --- | --- | --- | --- | --- |
| Cutpoint | Sensitivity | Specificity | Correctly classified | LR+ | LR- |
| ( >= 1 ) | 100.00% | 0.00% | 53.42% | 1.0000 |  |
| ( >= 2 ) | 87.18% | 35.29% | 63.01% | 1.3473 | 0.3632 |
| ( >= 3 ) | 87.18% | 82.35% | 84.93% | 4.9402 | 0.1557 |
| ( >= 4 ) | 82.05% | 82.35% | 82.19% | 4.6496 | 0.2179 |
| ( >= 5 ) | 82.05% | 88.24% | 84.93% | 6.9744 | 0.2034 |
| ( >= 6 ) | 76.92% | 94.12% | 84.93% | 13.0769 | 0.2452 |
| ( >= 7 ) | 74.36% | 97.06% | 84.93% | 25.2820 | 0.2642 |
| ( >= 8 ) | 71.79% | 97.06% | 83.56% | 24.4103 | 0.2906 |
| ( >= 9 ) | 71.79% | 100.00% | 84.93% |  | 0.2821 |
| ( >= 10 ) | 66.67% | 100.00% | 82.19% |  | 0.3333 |
| ( >= 11 ) | 53.85% | 100.00% | 75.34% |  | 0.4615 |
| ( >= 12 ) | 48.72% | 100.00% | 72.60% |  | 0.5128 |
| ( >= 13 ) | 33.33% | 100.00% | 64.38% |  | 0.6667 |
| ( >= 14 ) | 23.08% | 100.00% | 58.90% |  | 0.7692 |
| ( > 14 ) | 0.00% | 100.00% | 46.58% |  | 1.0000 |

| Obs | ROC area | Std. err. | Asymptotic normal<br>[95% conf. interval] |  |
| --- | --- | --- | --- | --- |
| 73 | 0.8790 | 0.0453 | 0.79027 | 0.96765 |

### 2) Further examples of epitope profiles of patient IgE vs. IgG4

In supplementary S-Fig. 1A to 1C, several serum examples of patients with ImmunoCAP class 3 to class 5 were shown in both IgE and IgG4 profiles with clinical data and data obtained by the 14 epitopes-ALL analyses.

**Supplementary S-Fig. 1A, 1B and 1C.**
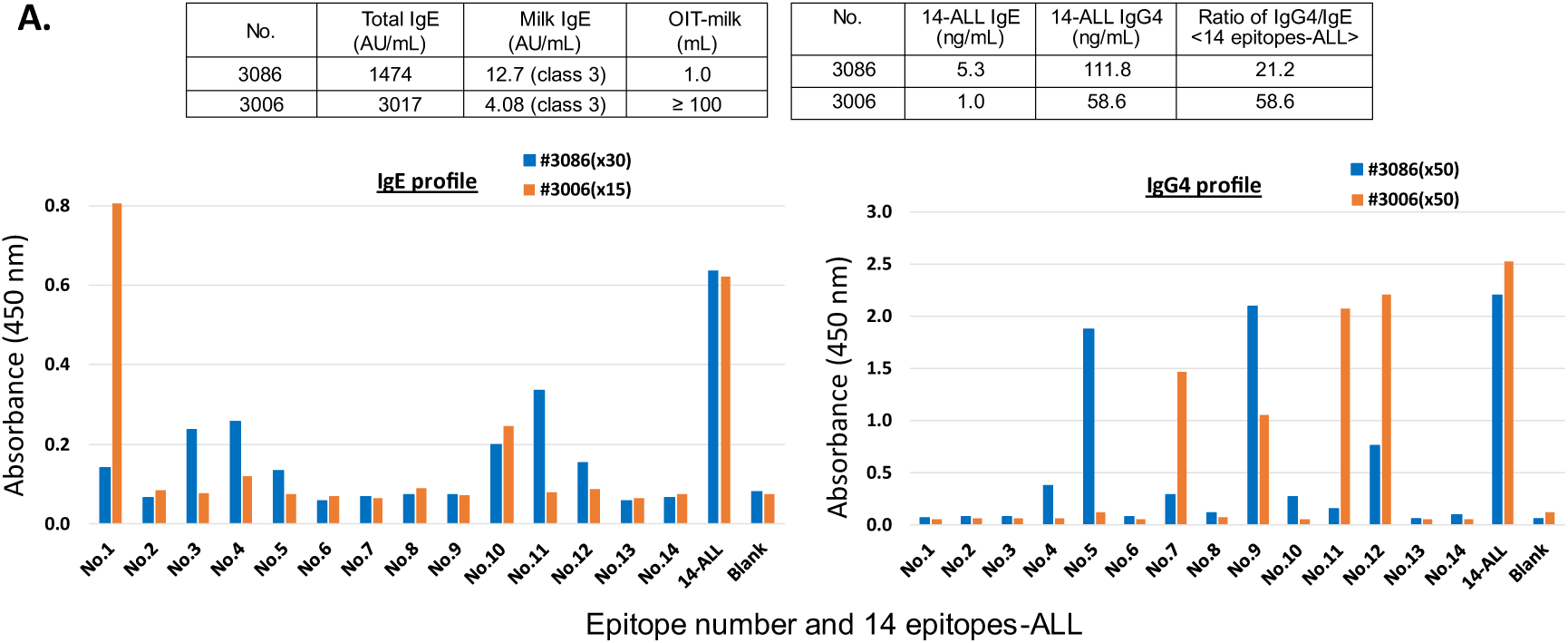
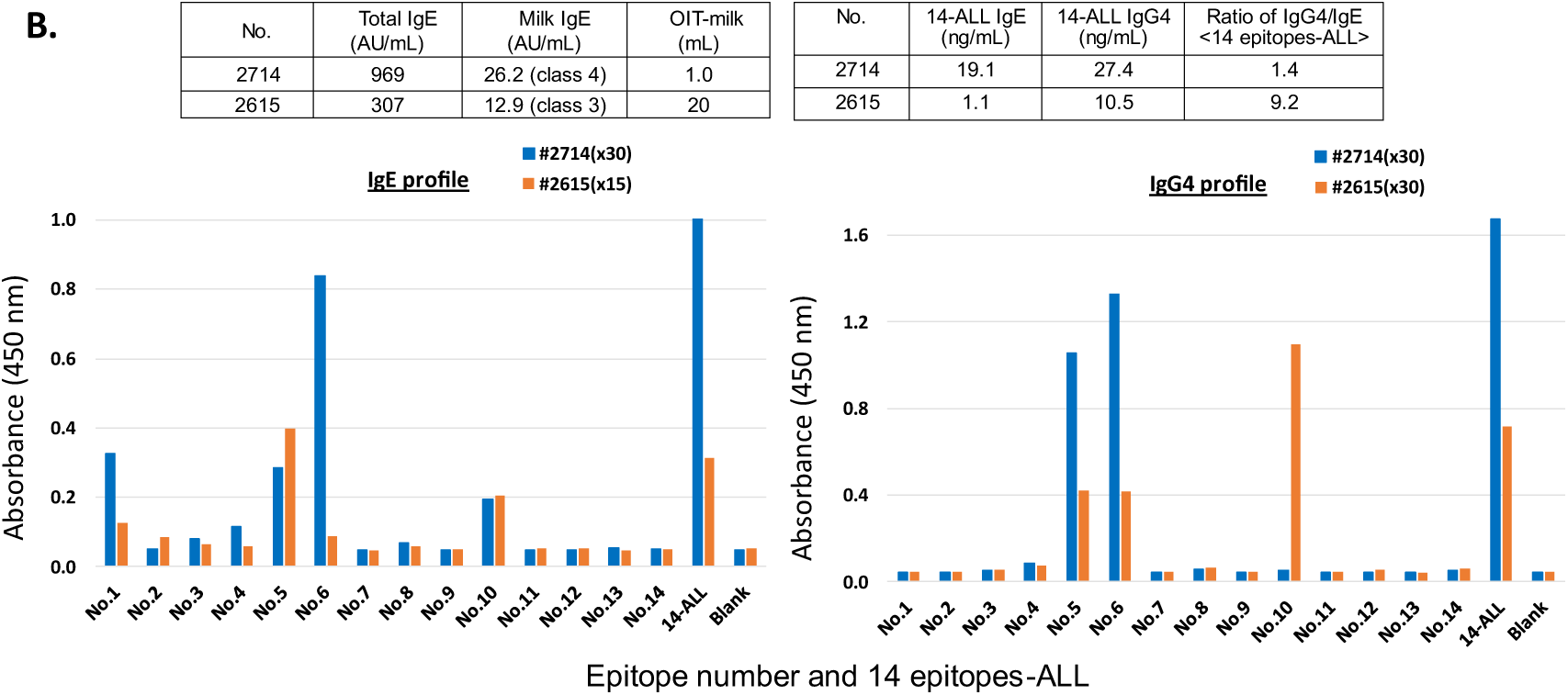
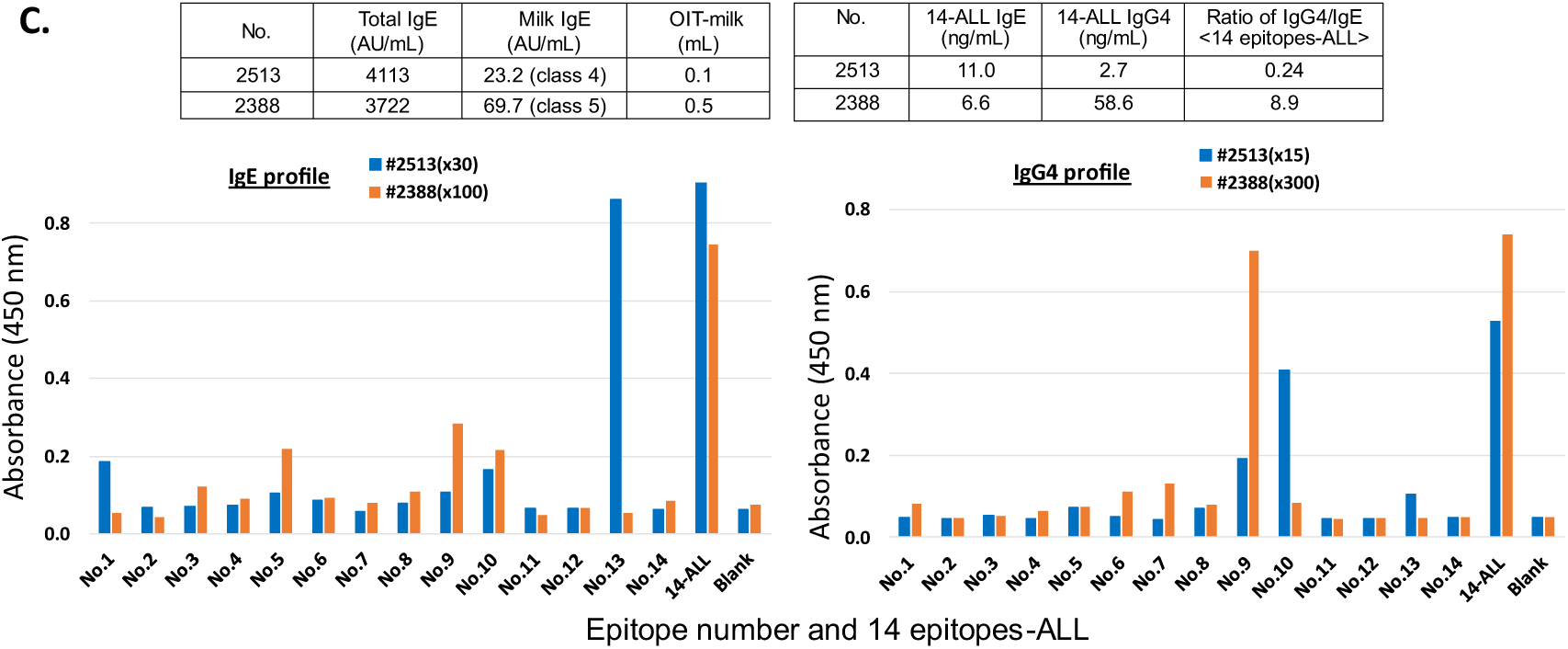
Additional profiles of IgE vs. IgG4 Abs. **A, B, C.** Further 6 examples of IgE/IgG4 profiles were shown. These are the profiles of imbalance between IgE vs. IgG4 (**A**), profiles possessing major epitopes in alpha Casein (**B**) and profile possessing rare pathogenic IgE-epitope (**C**). The clinical data such as total IgE and specific IgE detected by ImmunoCAP method as well as allowed volume of milk intake were listed in upper left table. In the upper right table, the IgE and IgG4 contents detected by 14 epitopes-ALL were also described.

In S-Fig. 1A, it was shown that patient-3006 was another example to demonstrate that No.1 epitope is nonpathogenic IgE epitope. In case of patient-3086, although IgG4 specific to No.5 and No.9 were largely induced, IgE Abs-recognizing No.11, No.4 and No.3 epitopes still exist and likely cause low amount of milk intake (1.0 mL). Regarding the IgE contents to the 14 epitopes-ALL, patient-3086 was over the cutoff criteria and patient-3006 was under the criteria.

It was further shown in S-Fig. 1B that patient-2714 had pathogenic No.6 epitope-specific IgE and low levels of pathogenic No.5 epitope-IgE. And large amounts of IgG4 specific to No.5 and No.6 were detected in right profile. This comparative analysis strongly suggested that IgG4 well compete No.5 epitope-specific IgE but that No.6- specific IgE did not reach sufficiently high levels to be blocked by the same epitope-recognizing IgG4 yet.

In the case of patient-2615, the IgE class by ImmunoCAP was 3, but the IgE content detected by 14 epitopes-ALL was low level at 1.1 ng/mL. Moreover, large amounts of No.5/No.6/No.10-specific IgG4 Abs were observed, resulting in the intake of 20 mL of milk. This profiling analysis indicates that this patient will soon be free from the limitation of milk intake when No.5-IgE Abs disappears.

A third other example is shown in S-Fig. 1C. In patient-2513, a low frequency IgE epitope of No.13 from beta Lactoglobulin (βLG) was observed, which is highly probable to be pathogenic, and very few IgG4 Abs specific to same No.13 and low level of No.10- recognizing IgG4 was observed. This patient was definitively responsive to beta LG as an allergen of cow’s milk. Thus, beta LG-containing whey milk is needed to increase IgG4 levels specific to the No.13 epitope as an OIT treatment.

In patient-2388, the IgE contents detected by 14 epitopes-ALL were over the cutoff criteria that is same with patient-2513, and No.5, No.9 and No.10 epitopes speific IgE were detected. Regarding IgG4 profile, No.9 epitope specific IgG4 was high but No.5 and No.10 epitopes specific IgG4 were not induced yet. This imbalance could be a reason why this patient can not increase milk intake while large amount IgG4 Abs exist over IgE content (IgG4/IgE : 8.9).

### 3) Inhibition of αCasein-IgE binding of sera pooled with OIT-resistant patients by the addition of OIT effective patient’s serum

In the earlier stage of this study, we used eight epitopes to preliminarily clarify IgE and IgG4 profiles for OIT-effective and OIT-ineffective patients. One OIT-effective patient with CMA (●101) had been able to drink milk 60 mL a day, resulting in very few IgE signals to casein. Significantly high IgG4 Abs were detected with only a positive No.5 epitope but not IgE responses in ●101 patient (Supplementary S-Fig. 2A and 2B). Moreover, this IgE-negative/IgG4-positive sera can inhibit IgE binding to αCasein in patient sera pooled with serum samples from 7 patients with severe CMA (Supplementary S-Fig. 2).

**Supplementary S-Fig. 2A, 2B and 2C.**
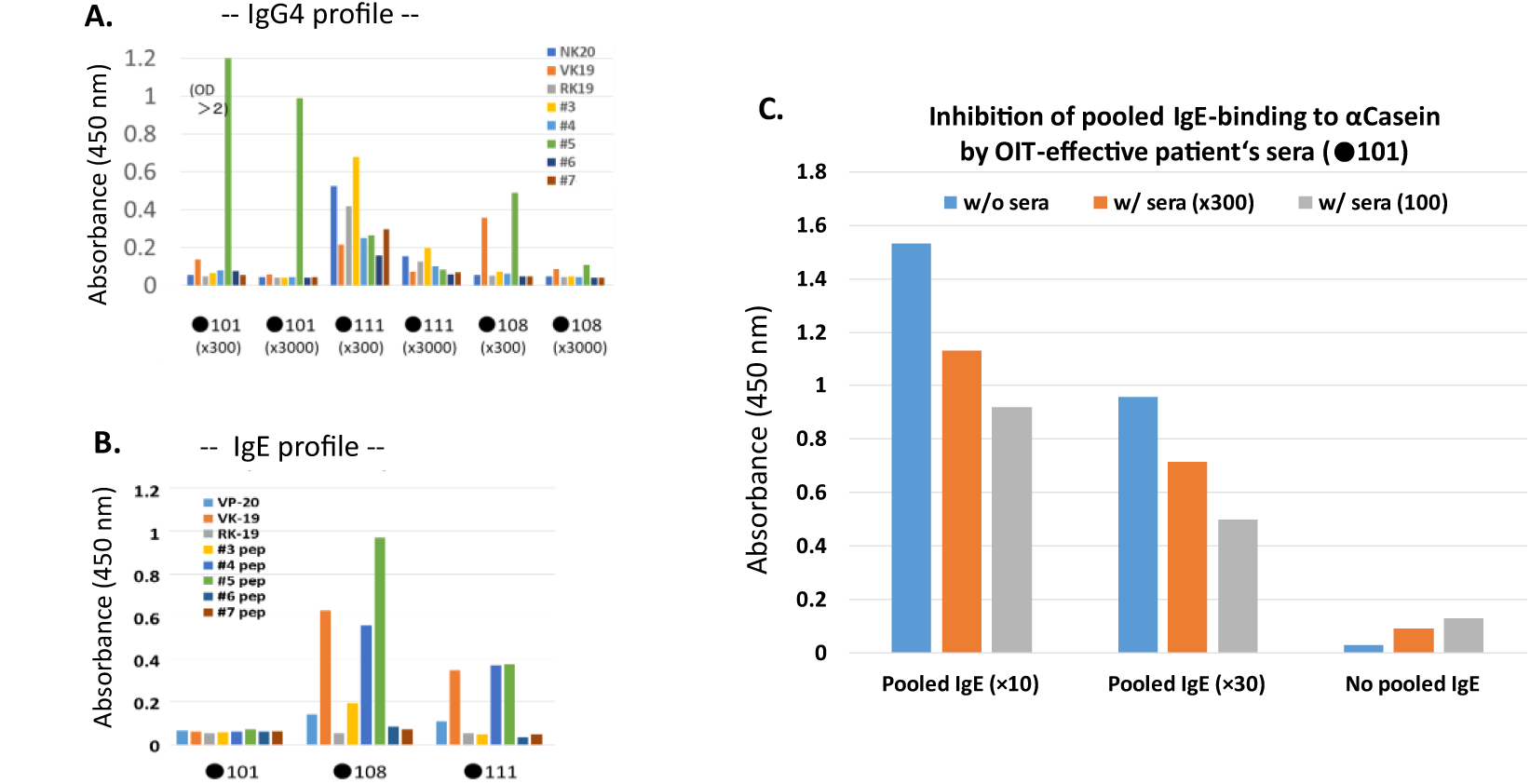
The patient serum skewed to one epitope-specific IgG4 without IgE can inhibit IgE responses recognizing αCasein. **A and B.** The eight epitopes were set to demonstrate IgE (**A**) and IgG4 (**B**) profiles in three serum samples of CMA patients had carried out oral immunotherapy of cow’s milk. The OIT-effective patent (●101)-derived serum sample had already skewed to one epitope-specific IgG4 (**A**), and there was no or few IgE Abs recognizing eight epitopes (**B**). **C.** The pooled sera of 7 patients with OIT-ineffective were inhibited their IgE binding to αCasein by addition of ●101 patient serum in the dilution-dependent manner. As shown in bar graph of right, IgE detection was very few signals that IgE content had already been very low. Left two bar graphs showed dilution dependent inhibition by adding ●101 serum samples in the IgE-αCasein binding.

### 4) Failure of correlation between IgG4 content or IgG4/IgE ratios vs, milk intake

In Fig. 2B and 2C, it has been shown that t the 14 epitopes-ALL may be applied for IgG4 as an indicator able to accurately estimate the blocking ability against IgE related to allergic reactions in patients with ImmunoCAP class 3. Although the previous publications have reported that specific IgG4 values detected by ImmunoCAP(Milk) did not correlated well with the activity of “blocking Abs” induced by OIT. (ref; 10-13), the data of Fig. 2B and 2C suggest that 14 epitopes-ALL is superior to allergen proteins of CMA in case of IgE detection. Therefore, we have further examined if there is any correlation of IgG4 contents and IgG4/IgE ratios specific to 14 epitopes-ALL vs. milk intake volumes obtained by OFC test.

All 38 serum samples were measured in terms of IgG4 content detected by the 14 epitopes-ALL; these IgG4 contents were used to calculate ratios of IgG4/IgE, using the previous IgE data shown in Fig. 2A (ref. 22). As shown in S-Fig. 3A and 3B, there was no obvious correlation between IgG4 contents or IgG4/IgE ratios vs. milk intake without allergic reactions. Even though the 14 epitopes-ALL can be useful for IgE monitoring, this is not the case in the contents of IgG4 specific to 14 epitopes-ALL, which were induced by OIT-related therapy of both OIT treatment and diet therapy of cow’s milk without eliciting allergic symptoms.

**Supplementary S-Fig. 3.**
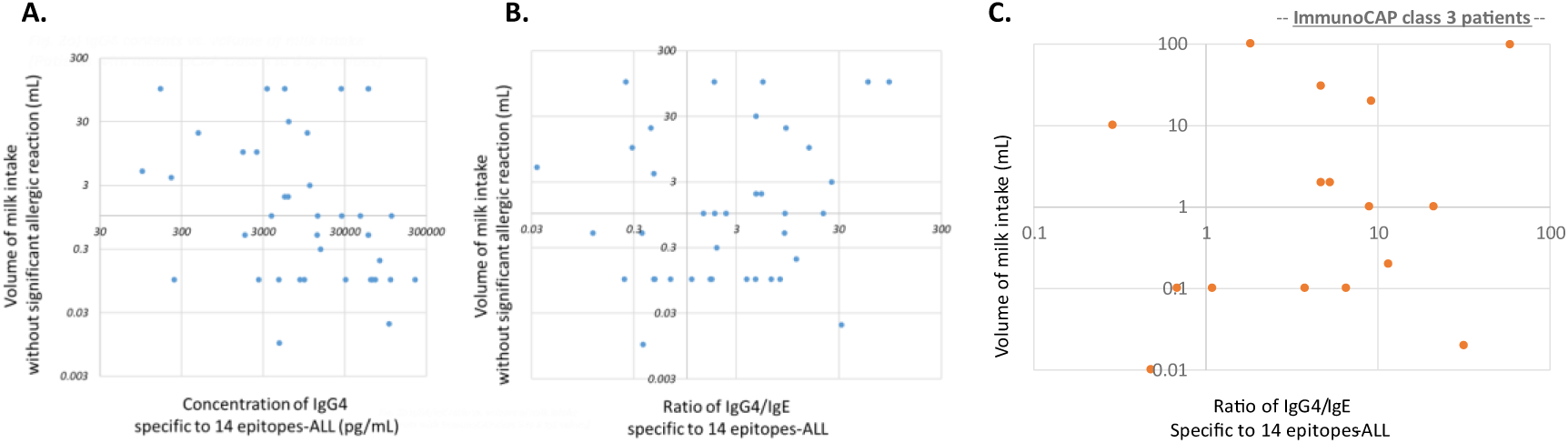
Trials of IgG4-content detected by the 14 epitopes-ALL as an indicator of IgE-blocking ability. **A and B.** Failure of relationship between allowed milk intake (volume) vs. IgG4 contents (**A**) or IgG4/IgE ratios (**B**). **C.** In the population of ImmunoCAP class 3 patients, there is also no correlation between milk intake vs. IgG4/IgE ratios.

These observations were similar to those of previous reports that have clarified no strong correlation between IgG4 and OIT effectiveness (ref. 10-14).

## Notes

### Competing Interest Statement

The authors have declared no competing interest.

### Author Declarations

The study was approved by the University of Fukui Ethics Committee. Non-CMA volunteers at Kanazawa University Hospital also participated in this study (No. 2024-148, No. 2024-131), which was approved by the Kanazawa University Ethics Committee. Informed consent was obtained in accordance with the principles of the Declaration of Helsinki.

